# Applications of Digital Microscopy and Densely Connected Convolutional Neural Networks for Automated Quantitation of Babesia-Infected Erythrocytes

**DOI:** 10.1101/2021.04.27.21256115

**Authors:** Thomas JS Durant, Sarah Dudgeon, Jacob McPadden, Anisia Simpson, Nathan Price, Wade Schulz, Richard Torres, Eben M Olson

## Abstract

**Background:** Clinical babesiosis is diagnosed, and parasite burden is determined, by microscopic inspection of a thick or thin Giemsa-stained peripheral blood smear. However, quantitative analysis by manual microscopy is subject to observer bias, slide distribution errors, statistical sampling error, recording errors, and is inherently burdensome from time management and workflow efficiency standpoints. As such, methods for the automated measurement of percent parasitemia in digital microscopic images of peripheral blood smears could improve clinical accuracy, relative to the predicate method.

**Methods:** Individual erythrocyte images (shape: 70×70×3) were manually labeled as “parasite” or “normal” and were used to train a model for binary image classification. The best model was then used to calculate percent parasitemia from a clinical validation dataset, and values were compared to a clinical reference value. Lastly, model interpretability was examined using an integrated gradient to identify pixels most likely to influence classification decisions.

**Results:** The precision and recall of the model during development testing were 0.92 and 1.00, respectively. In clinical validation, the model returned increasing positive signal with increasing mean reference value. However, there were two highly erroneous false positive values returned by the model. Lastly, the model incorrectly assessed three cases well above the clinical threshold of 10%. The integrated gradient suggested potential sources of false positives including rouleaux formations, cell boundaries, and precipitate as deterministic factors in negative erythrocyte images.

**Conclusions:** While the model demonstrated highly accurate single cell classification and correctly assessed most slides, several false positives were highly incorrect. This project highlights the need for integrated testing of ML-based models, even when models in the development phase perform well.

## INTRODUCTION

Clinical Babesiosis is a haemoprotozoan disease that is most commonly transmitted from animals to humans by invertebrate vectors (e.g., *Ixodes scapularis*, the black legged deer tick)(1). In the United States, 95% of cases occur in the Northeast and Upper Midwest states, occurring primarily between May and October. In the state of Connecticut, the seroprevalence has been shown to range between 0.3-17.8%, with the number of reported cases being approximately 44 per 100,000 (2). Disease severity can range from asymptomatic to severe, the latter of which may lead to life-threatening scenarios. Severe disease is more common in specific at-risk populations including those who are post-splenectomy, immunocompromised, or older than 50 years of age. The all-cause mortality of babesiosis has been estimated as <1% for clinical cases, and approximately 10% for iatrogenic cases (e.g., transfusion-transmitted) (2).

The diagnostic gold standard for babesiosis is microscopic inspection of thick, or thin, Giemsa-stained peripheral blood smear (1). If *Babesia* spp is identified, the degree of parasitemia is used to guide patient management strategies. For mild disease, or minimal parasitemia, antimicrobials are the preferred therapy. However, the American Society for Apheresis (ASFA) guidelines state that severe babesiosis is a category II indication for red blood cell (RBC) exchange. Severe disease is determined both by clinical and laboratory criteria including significant parasitemia (e.g., >10%), the presence of comorbidities (e.g., asplenia), or severe symptoms such as, disseminated intravascular coagulation or multiorgan failure (2). While there is no consensus on when to discontinue RBC exchange, it is recommended that patients with severe babesiosis be monitored closely, with parasitized erythrocytes quantified daily alongside continued RBC exchange until parasite burden decreases below 5% (2,3).

Percent parasitemia is the quotient of parasite-infected erythrocytes over the number of total erythrocytes counted. To derive this in a clinical laboratory, the process commonly involves a medical laboratory scientist (MLS) counting a large number of erythrocytes (e.g., 1,000) using a 100x oil-immersion objective. While this process requires minimal laboratory equipment, it does require an experienced MLS to ensure optimal accuracy and reproducibility for serial measurement purposes (1). In addition, quantitative analysis by manual microscopy is subject to observer bias, slide distribution errors, statistical sampling error and recording errors, and is inherently burdensome from time management and workflow efficiency standpoints (4,5). Such limitations can mislead or delay therapeutic decision making, particularly in the context of therapeutic RBC exchange. Accordingly, there remains a significant need to develop automated methods to optimize the cost, efficiency, and accuracy of quantitative analysis.

The progress made in computer vision and machine learning (ML) technology over the last decade has encouraged a corresponding increase in their implementation in the clinical laboratory (6). With the decreasing availability of experienced medical laboratory scientists, evaluating ML-based software capabilities without expert operator review remains an important consideration in study design (7,8). To this end, we sought to develop and evaluate the accuracy of a an ML-based method for the automated measurement of percent parasitemia in digital microscopic images of peripheral blood smears. Specifically, we sought to describe the accuracy of parasitemia measurements, as determined by ML-based software, relative to an MLS-derived reference standard (MLS-RS). We hypothesized that results generated by the ML-based software would show superior precision to MLS-RS while achieving clinically comparable numerical results to the average MLS-RS.

## METHODS

### Hardware and Operating Systems

Computation for model training was performed on a local Linux server (NVIDIA DGX Server Version 4.6.0) (GNU/Linux 4.15.0-122-generic x86_64) running Ubuntu (version: 18.04.5 LTS). Processing hardware included 80 CPUs (Intel(R) Xeon(R) CPU E5-2698 v4 @ 2.20GHz) and 8 GPUs (Tesla V100-SXM2-16GB) using CUDA Toolkit (version: 11.0).

### Data Set Curation

This study has been reviewed and approved by the Yale University Internal Review Board (IRB# 2000020244). Clinical blood samples were originally collected as part of routine clinical workflow in lavender-top (EDTA) tubes for screen and quantitation of *Babesia* spp. Slides and concomitant digital images of the associated peripheral blood smears, which were found by to be positive for *Babesia* spp and negative for *Malaria* spp (BinaxNOW Malaria; Abbott, Chicago, IL), were flagged for inclusion using previously described methods (9–11). Slides and the concomitant digital images of *Babesia*-negative samples were collected from the routine clinical workflow throughout the study period and reviewed by a clinical pathologist for the absence of *Babesia* spp prior to inclusion.

Slides and images were separated into two distinct groups, representing separate patient cohorts: (1) The model development dataset and (2) the clinical validation dataset. The model development dataset was used for training, validation, and preliminary evaluation of the cell classification model. The clinical validation dataset was used as a second, ‘external’ validation dataset to evaluate how the model would perform in a clinical implementation workflow, as compared to a predicate method-based reference standard.

All peripheral blood smears were created and imaged on a DI-60 Integrated Slide Processing System (Cellavision AB, Lund, Sweden). The DI-60 uses a 100X-objective and a 0.5X magnifier prior to imaging, rendering an effective magnification of 50X. Images are 3-channel RGB, with a resolution of 5 pixels per micron. In the model development dataset, slide images had an average height and width of 2884 pixels (95% CI: 2882-2885) and 2867 pixels (95% CI: 2865-2868) (Figure 1A). Slides included in the model development dataset were imaged a single time. Slides included in the clinical validation dataset were imaged three times on the same scanner to compute intra-precision for quantitation of *Babesia* spp during subsequent portions of the study.

**Figure 1:**
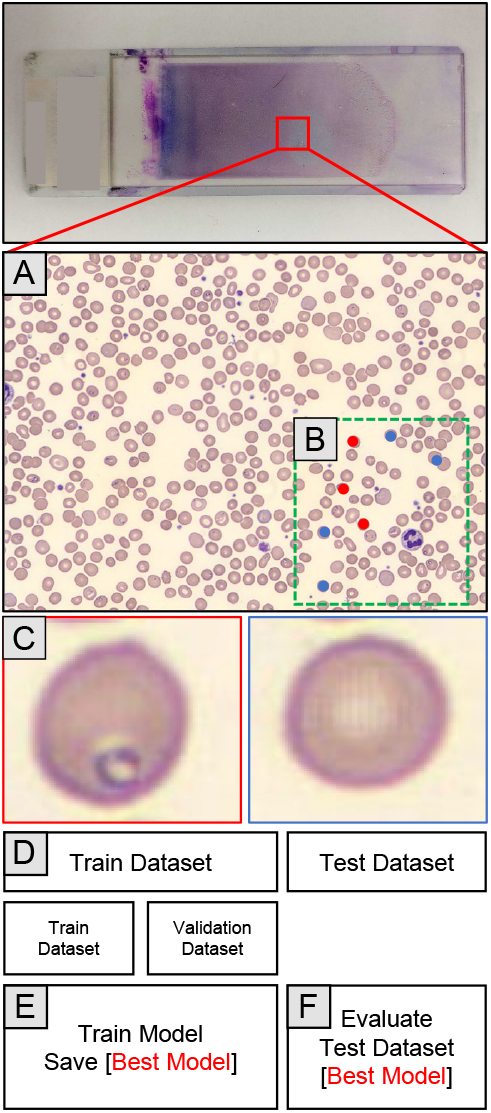
Flow diagram of model development process. (A) Slides included in the model development dataset were imaged a single time by the Cellavision DI-60 and uploaded to a custom-built-web application for label annotation. (B) Central X-Y coordinates of infected (red) and non-infected (blue) erythrocytes were marked on the slide-level images. (C) Central X-Y coordinates were used to crop individual erythrocytes into 70 × 70 pixel, 3-channel arrays and paired with the corresponding label of either ‘parasite’ (red) or ‘normal’ (blue). (D) Labeled erythrocyte images were collectively divided 80:20 into train and test datasets, respectively. The train dataset was further subdivided 70:30 into train and validation datasets, respectively. (E) The train and validation datasets were used train the image classification model. (F) Following completion of training, the [best model] was used to evaluate model performance using the test dataset.

### Cell Labeling for Model Development Dataset

Slide-level images from the model development dataset were uploaded to a custom-built web application for labeling of individual erythrocytes using one of two labels: (1) parasite or (2) normal. Using the web application, annotators marked central X-Y coordinates of infected and non-infected erythrocytes (Figure 1B). X-Y coordinates of cell centers were then used to crop individual erythrocytes from the slide-level parent image into 70×70 pixel, 3-channel image arrays. These 70×70×3 images were then paired with their corresponding label of either ‘parasite’ or ‘normal’ (Figure 1C). The labeling process was performed by a single laboratory medicine attending and author of this manuscript (TJD). As a post-processing step, X-Y coordinates which were within 140 pixels of another set of X-Y coordinates were removed from the dataset following completion of the annotation process. This was done to ensure that there was no overlap of images in the final development dataset which, if present, could have resulted in part of an image being represented in both the training and validation and test datasets, leading to overfitting, or an over-optimistic estimate of model performance.

Ultimately, the final dataset used for model development consisted of non-overlapping, individual erythrocyte images (shape: 70×70×3) with an associated label of ‘parasite’ or ‘normal’. These data were split and used to train, validate, and test the image classification model. The model development dataset was divided 80:20 into train and test datasets, respectively (Figure 1D). The train dataset was further subdivided 70:30 into train and validation datasets, respectively. The train and validation datasets were used during the training of the image classification model (Figure 1E). The test dataset was used to evaluate model performance following completion of training (Figure 1F).

### Network Implementation

For image classification, the authors implemented DenseNet121 as the base model, initialized with pretrained weights from ImageNet (7). Densely connected neural networks were first described by Huang et al. and are a commonly used architecture for learning image classification tasks (8). This neural network was chosen based on previously published performance metrics comparable with current state of the art models, and because it uses relatively fewer parameters, making it faster to train and easily portable (12). Base model layers were not frozen and were configured as trainable. DenseNet121 was combined with a custom set of prediction layers, specific to this image classification task. These included a 2-dimensional global average pooling layer, a dropout layer, and a densely connected layer with sigmoid activation function for binary classification. The Adam method was used for gradient-based optimization. In total, there were 7,038,529 parameters, 6,954,881 of which were trainable. The network was implemented using Tensorflow (version: 2.4.0rc0), Tensorflow-gpu (version: 2.4.0rc0) and Python (version: 3.6.9).

### Model Development Protocol

Train dataset images were subjected to label preserving augmentation prior to being served as input to the network. Image augmentation included random horizontal and vertical flips, random rotation, random translation, random zoom, random contrast adjustments, and random brightness adjustments. Lastly, due to the imbalanced nature of our training dataset the ‘parasite’ class was oversampled to produce a 1:1 ratio of parasite and normal images during training. The network was trained for a total of 50 epochs (i.e., iterations) over the complete training dataset. The validation dataset was used to monitor model performance during training for subsequent tuning according to the calculated binary cross-entropy loss. Model parameters were saved following a reduction in the binary cross-entropy loss, calculated from the validation dataset after each epoch. The initial learning rate was set to 1e-5 and decreased by a factor of 10 if validation loss did not improve after 5 epochs. The total training process was repeated three times using unique random seed initializers to evaluate variability in train performance metrics. Performance metrics monitored during training included true positives (TP), false positives (FP), true negatives (TN), false negatives (FN), binary accuracy, precision (i.e., positive predictive value) (TP / (TP + FP)), recall (i.e., sensitivity) (TP / (TP + FN), and area under the receiver operator characteristic curve (AUC). These were calculated on both train and validation datasets following the completion of each epoch. Following model training, the best model parameters (i.e., those which achieved the lowest validation loss) were used to evaluate individually labeled erythrocytes in the test dataset. Cells with a probability score greater than or equal to 0.5 were assigned ‘parasite’ prediction labels. Test predictions were then used to calculate the performance metrics for the test dataset. Similarly, the ‘best model’ was used to evaluate cells in the clinical validation protocol.

### Clinical Validation Protocol

Following model development, a separate set of peripheral blood smear slides were used to assess the accuracy of the model in a simulated clinical workflow. Due to the inherent variability seen with quantitative analysis by microscopy, a clinical reference standard consisting of multiple measurements was compiled for comparisons between the model and the predicate method. Accordingly, each glass slide in the clinical validation dataset was independently evaluated by three MLS’s with 26, 6, and 4 years of experience for MLS A, B, and C, respectively. The clinical validation slides were shuffled, specimen numbers on the glass slides were covered, and a box containing the clinical validations slides was given to each of the MLS’ for independent evaluation. Each MLS evaluated all clinical validation slides three separate times (Figure 2A). In total, this process generated 9 results of percent parasitemia for each slide in the clinical validation dataset. These data were used to calculate the average percent parasitemia across all 9 reads which was used as the MLS-RS for each case/sample (Figure 2B). Of note, the lower limit of quantitation for percent parasitemia in the clinical laboratory at our institution is 1% and results below this value are reported out as <1% in routine practice. For the purposes of this study, MLSs were asked to record the precise parasitemia value, including those below 1%, to allow for a completely empirical comparison against the model.

**Figure 2:**
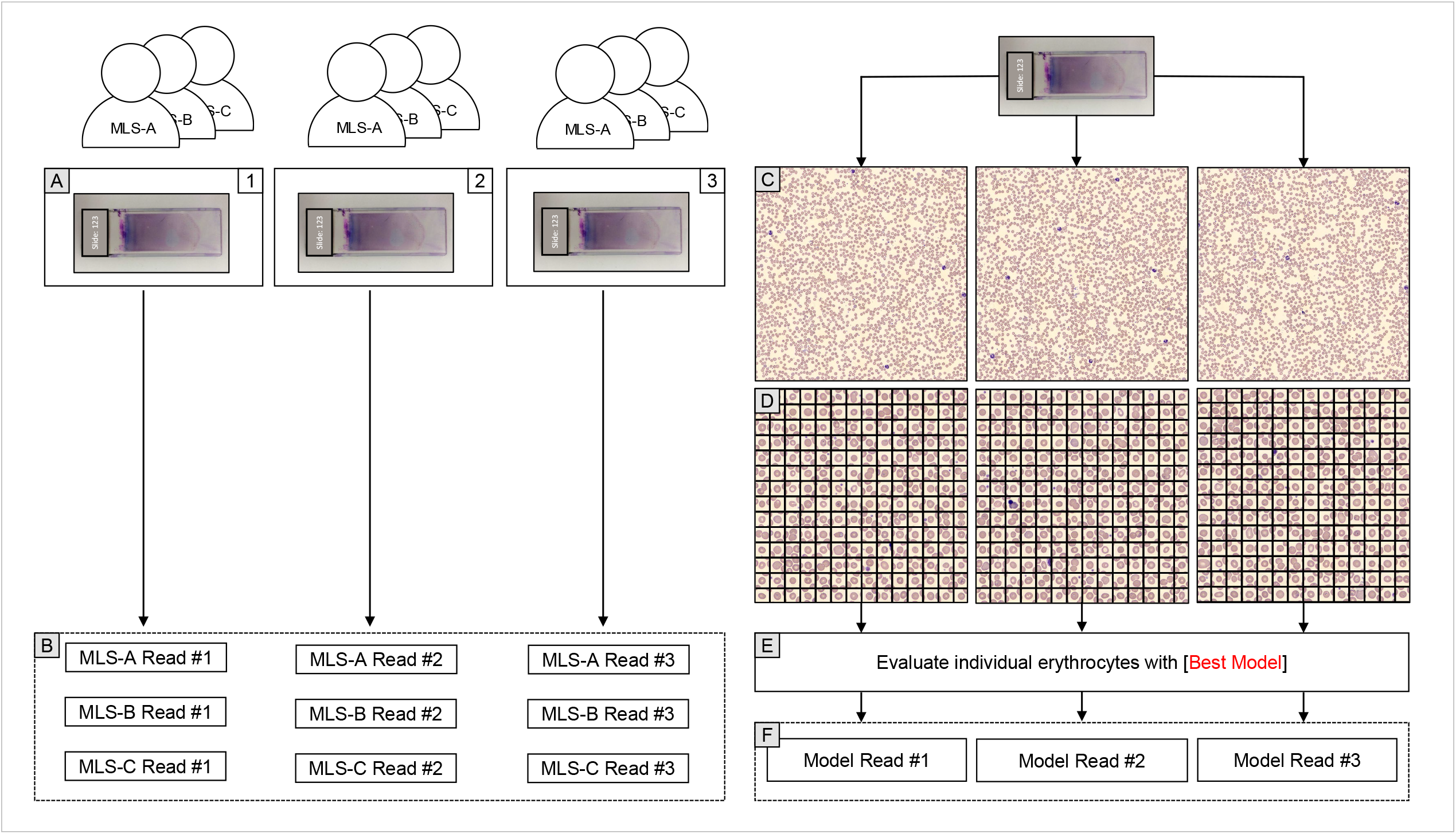
Flow diagram of clinical validation process. (A) Each peripheral blood smear was evaluated three times, in a blinded fashion, by each MLS. (B) This process yielded a total of 9 parasitemia results for each slide in the clinical validation dataset. These data were used to calculate the average parasitemia across all 9 reads which was used as the clinical reference standard for each case. (C) Each glass slide in the clinical validation dataset was imaged three separate times by the Cellavision DI-60. (D) Contour-based cell segmentation was used to extract individual erythrocytes from the DI-60 slide-level images as 70×70×3 cropped images. (E) Individually cropped erythrocytes were independently evaluated by the [best model] to yield a predicted class (i.e., ‘parasite’ or ‘normal’). (F) The number of cells with the predicted label of ‘parasite’ were divided by number of total cells classified to yield the parasitemia result. This process was done one time for each DI-60 image. With three images per specimen, this yielded a total of 3 parasitemia results per slide, which were used to calculate an average parasitemia result for each specimen.

For the model-based method, as mentioned, each slide in the clinical validation dataset was scanned three separate times by the DI-60 (Figure 2C). A custom cell-segmentation script was then used to crop individual erythrocytes from the peripheral blood smear image (Figure 2D). Cell-segmentation was implemented using OpenCV (version: 4.2.0.34) using contour-based (cv.findContours()). Individual erythrocytes (shape: 70×70×3) were then provided as input to the best model, as defined in the development protocol, to yield a predicted class (i.e., ‘parasite’ or ‘normal’) for each individually cropped erythrocyte (Figure 2E). Following classification of individual erythrocytes, the number of cells with the predicted label of ‘parasite’ were divided by number of total cells classified to yield the quantification of percent parasitemia. This process was done one time for each image with three images per specimen, yielding a total of 3 parasitemia results per slide (Figure 2F).

Method-to-method (i.e., accuracy) comparisons between the model and MLS-RS percent parasitemia were made using a variety of approaches: (1) bar plot visualization; (2) regression and Bland-Altman plots; (3) quantitative agreement of model percent parasitemia in relation to ±2 SD of the average MLS-RS percent parasitemia (n=9) for each case in the clinical validation dataset; (4) categorical agreement of percent parasitemia bins; (5) categorical agreement around the clinical decision threshold of 10%. Precision was assessed using the coefficient of variation, which was calculated on a case-wise basis across the MLS (n=9) and model results (n=3).

### Model Interpretability

In an effort to examine the relationship between model predictions and image features, we implemented an explainable artificial intelligence (XAI) technique based on axiomatic attribution for deep networks and known as Integrated Gradients (IG) (13). While the methods of IG are outside the scope of this report, the general purpose is to identify pixels within each image which most heavily influence a model’s prediction, and derived from the gradient (i.e., slope or derivative) of the prediction function relative to each feature (i.e., pixel). For the purposes of this report we attempted to provide representative samples of what we observed when reviewing the images derived from an IG implementation. This was done on the test images in the model development dataset.

## RESULTS

### Dataset Curation

A total of 96 unique slides were included in this study. Of these, 71 slides were included in the development dataset, 28 of which were found to be positive for *Babesia* spp by routine clinical workflow. A total of 14,633 individual erythrocyte images were initially labeled. Of those, 2,019 images that had overlapping cells were removed, yielding a final development dataset of 11,388 erythrocytes labeled as normal and 1,226 with a parasite. The mean number of labeled cells per unique slide was 178 (SD 63; range 1-286). Of the slide-level images which were *Babesia-*positive, the mean parasitemia was 6.5% (SD 4.5; range 1.0-20.0). The clinical validation dataset consisted of the remaining 25 slides, of which 64% (n=16) were *Babesia*-positive. The mean parasitemia among the *Babesia*-positive slides in the clinical validation dataset was 8.9% (SD 9.4; range 1.0-29.2).

### Model Development

The cell classification model was trained 3 separate times. Each training replicate consisted of 50 epochs (iterations). Learning rates decayed following validation loss plateau across all training replicates, with the final value ranging from 1e-8 to 1e-9. Minimum validation loss was observed following completion of training epoch 22, 22, and 31 for each of the training replicates, with an average binary cross-entropy of 0.024 (SD 0.003). Binary cross-entropy loss was plotted and inspected for positive divergence of validation loss, relative to training loss, as an empirical indicator of overfitting. This was observed minimally in the later training epochs (Figure 3A). Precision, recall, and AUC for asymptotically approached model performance limits which were concordant with plateaus of validation loss, indicating model improvement to be unlikely to occur with additional training iterations (Figures 3B-D). Training replicate 3 achieved the lowest validation loss during training (0.021) and was subsequently used for evaluation of the test and clinical validation datasets. Model predictions on the test dataset resulted in 20 false positives and zero false negatives. The precision and recall were 0.92 and 1.00, respectively (Figure 4A). The binary classification accuracy was 0.99. The distribution of predicted probabilities for erythrocytes in the test dataset was visualized and demonstrated a predominantly bimodal distribution between the predicted classes (Figure 4B).

**Figure 3:**
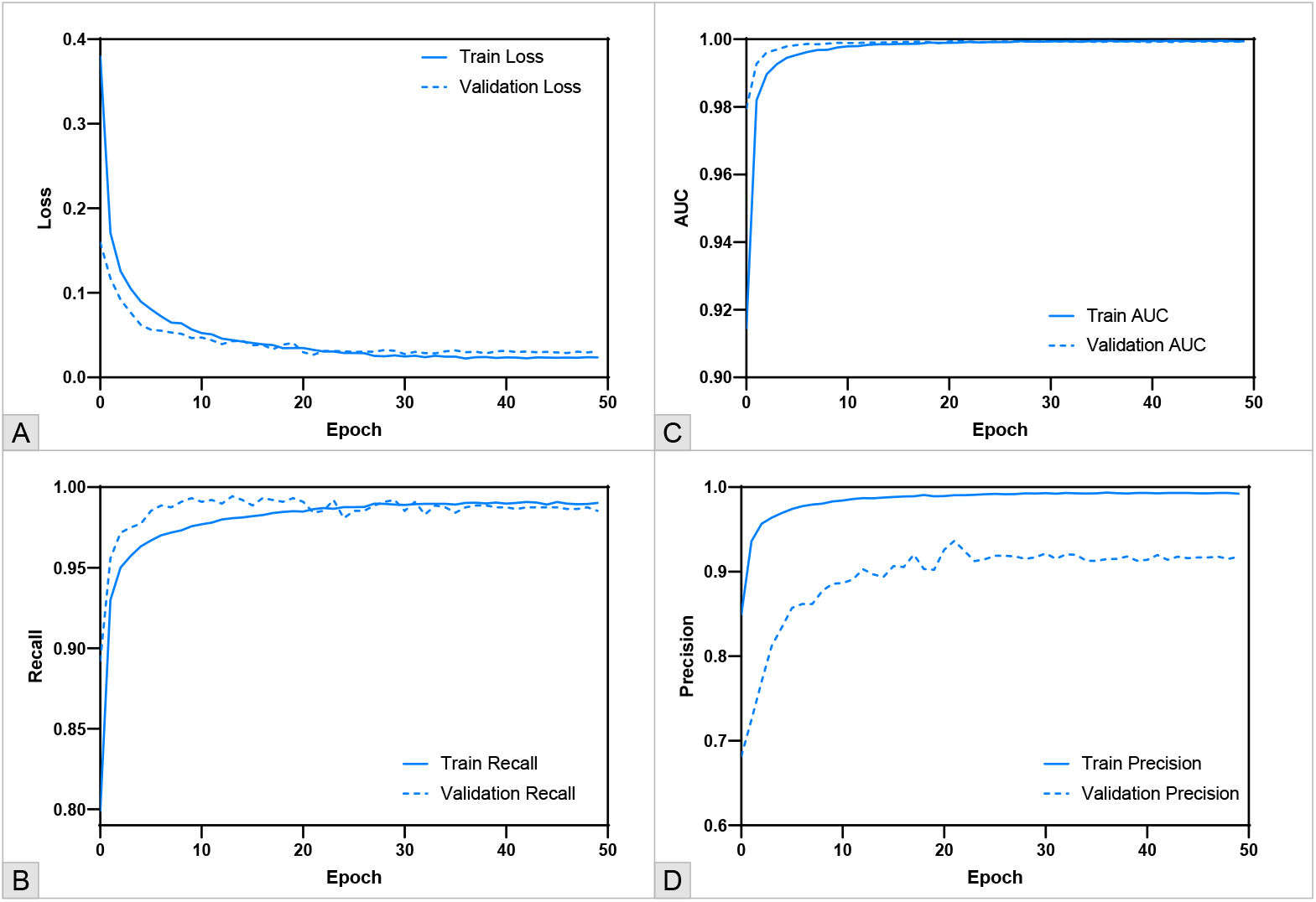
Model performance metrics plotted as a function of training epochs (iterations). (A) Train and validation loss. (B) Train and validation recall (sensitivity). (C) Train and validation area under the receiver operator characteristic curve. (D) Train and validation precision (positive predictive value).

**Figure 4:**
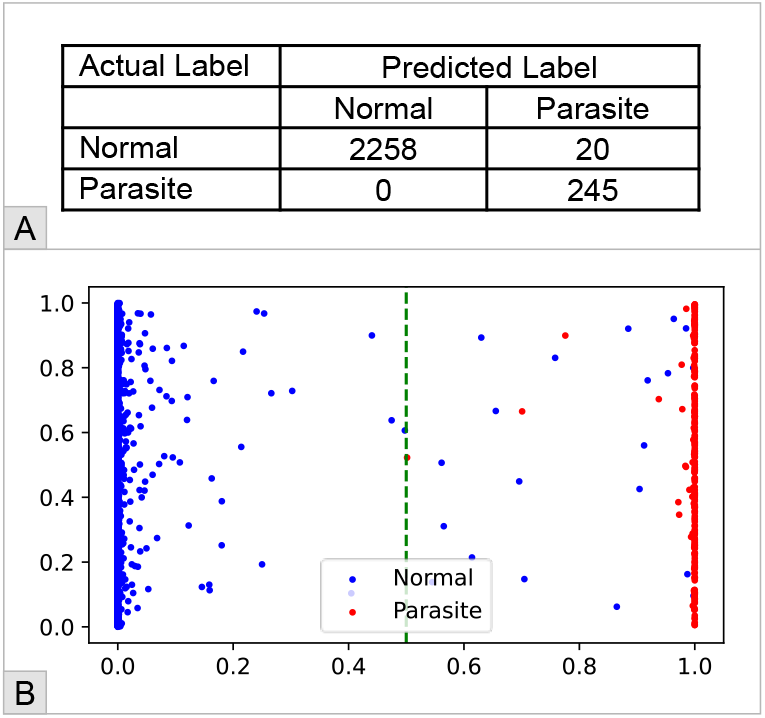
Model classification results on test dataset. (A) Confusion matrix of actual versus predicted labels. (B) Per-cell probability distribution of model predicted class with actual labels depicted in color (red = parasite) (blue = normal). X-axis: The probability of the predicted class being ‘parasite’. Y-axis: Random number between 0 and 1 was assigned to each cell for better visualizing data points. Green dotted line: Decision threshold for prediction label of ‘parasite’ – i.e., cells with a predicted probability of ≥ 0.5 are labeled as ‘parasite’.

### Clinical Validation of Model-Based Method

A total of 25 unique slides were identified for evaluation in the clinical validation set, 16 of which were found to be positive for *Babesia* spp by routine clinical workflow. Of those 16, one (Case #15) was excluded from analysis, as per the consensus recommendation of the participating MLS’ due to excessive artifact, Howell-Jolly bodies, and only rare, dying parasites. The remaining slides were evaluated in three separate instances by each of the MLS’ with an average parasitemia ranging from <0.1% to 38.5% (Supplemental Table 1 and Supplemental Figure 1).

Model classification demonstrated an increasing positive signal (i.e., higher parasite count) with respect to the MLS-RS; however, the automated model also demonstrated spurious positive signal with the negative cases (Cases 16-25). In addition, the model returned highly erroneous false positive signal on cases 11 and 16, relative to the MLS-RS (Figure 5). A simple linear regression was performed to evaluate the concordance between the MLS-RS and the model predictions. The regression equation was determined as: 4.78 + 0.55x with correlation coefficient (R^2^) of 0.244 (Figure 6A). With cases 11 and 16 removed, the regression equation is calculated as: 1.68 + 0.68x with an R^2^ of 0.916. Bland-Altman plots were also assessed for bias trends, and similarly demonstrate erroneous positive signal on the low end and erroneously low positive signal on the high end (Figure 6A and 6B).

**Figure 5:**
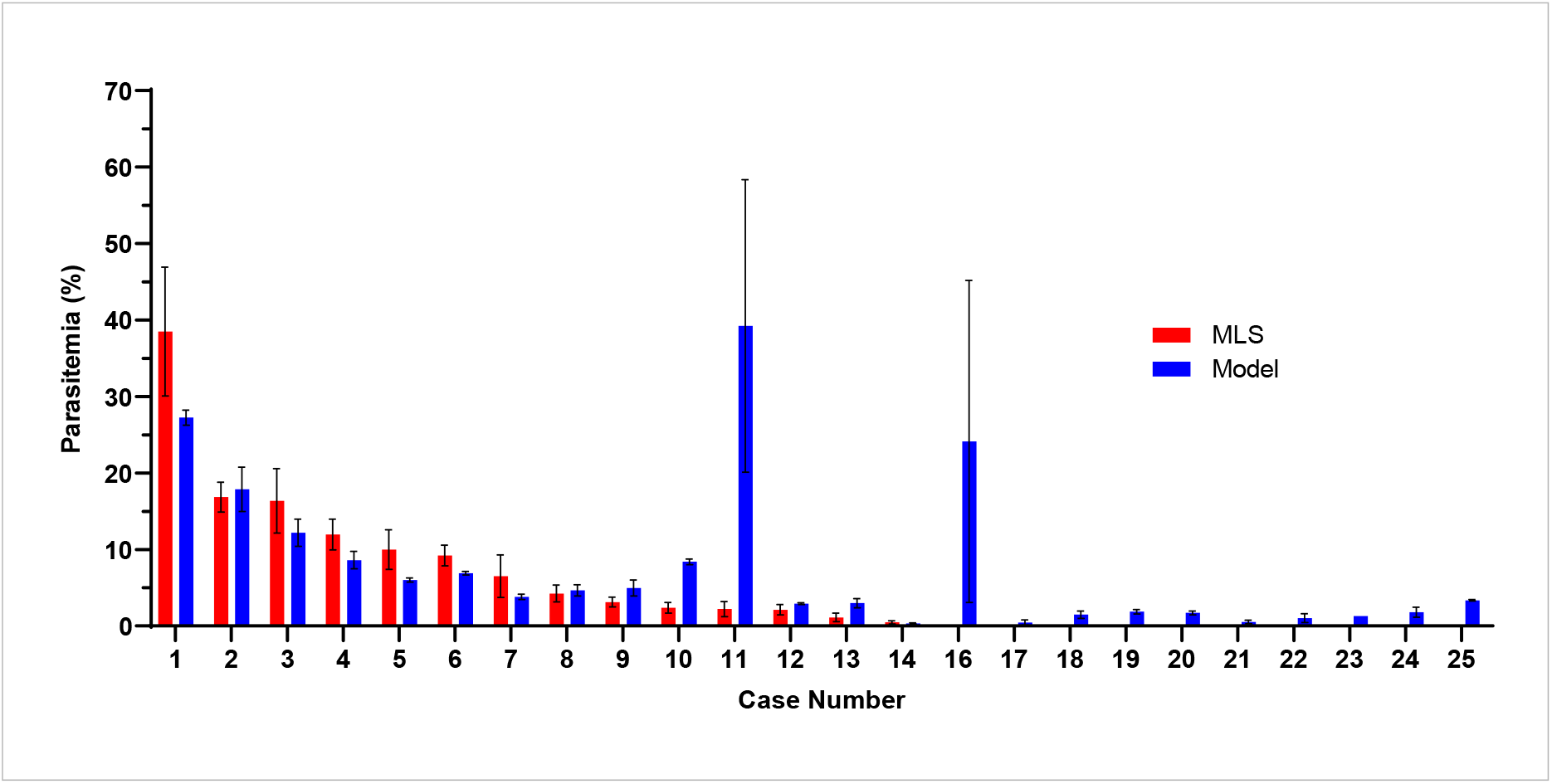
Bar plot of mean percent parasitemia for the MLS-RS (n=9) and the model-based method (n=3). Error bars represent 1 standard deviation.

**Figure 6:**
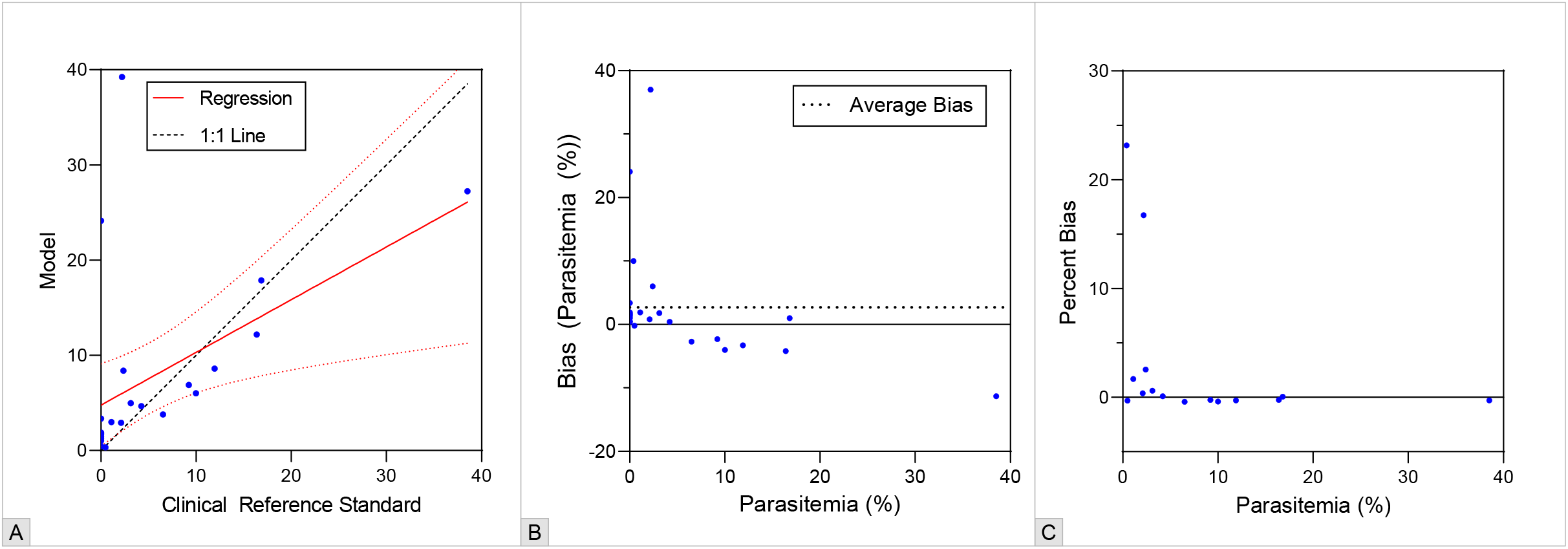
Visualizations for method-to-method comparison of MLS-RS and model-based method. (A) XY-scatter plot with regression line overlay (red-dotted line represents 95% confidence interval of regression). (B) Bland-Altman absolute bias plot. (C) Bland-Altman percent bias plot.

Of the 14 positive cases included in the clinical validation dataset, 10 were within 2 SD of the MLS-RS mean. However, only 7 were concordant between the model and MLS-RS with regards to the percent parasitemia bins. In addition, there were three major errors by the model-based method, which were defined as discordance around the clinical decision point of 10% parasitemia. Of the 14 positive cases, the MLS-RS CV was less than 20% in only 3 cases, whereas the Model CV was less than 20% for 10 of the cases (Supplemental Table 2).

### Model Interpretability

Cells from the test dataset and the clinical validation dataset were evaluated using the IG approach to visualize feature pixel-level activation patterns. Cells from the test dataset generally demonstrated activation of pixels which were near the intra-erythrocytic parasite (Figure 7). Cells from case 25, a negative case in the clinical validation set, were also examined and demonstrated erroneous activation on non-parasitic features. Some of these features included erythrocyte abnormalities (e.g., target cell contours), precipitate, and overlying platelets. In some cases, the model appeared to be focusing on background pixels which may be indicative of overfitting in some aspects of the model (Figure 8).

**Figure 7:**
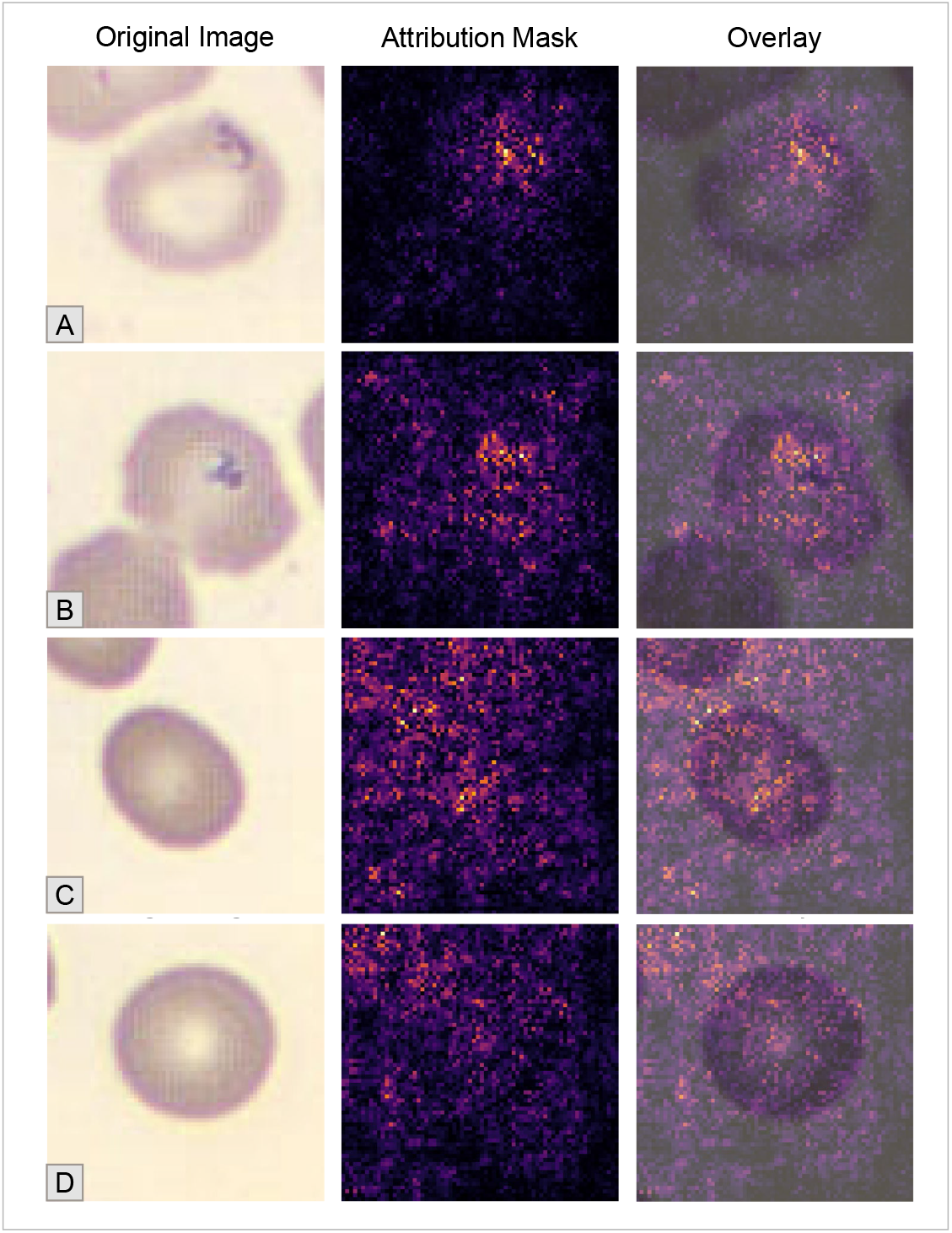
Integrated gradient (IG) visualizations including the original image, the pixel-wise IG attribution mask, and the overlay of the two. Images are from the model development test dataset. (A and B) Representative examples from the ‘parasite’ class. (C and D) Representative examples from the ‘normal’ class.

**Figure 8:**
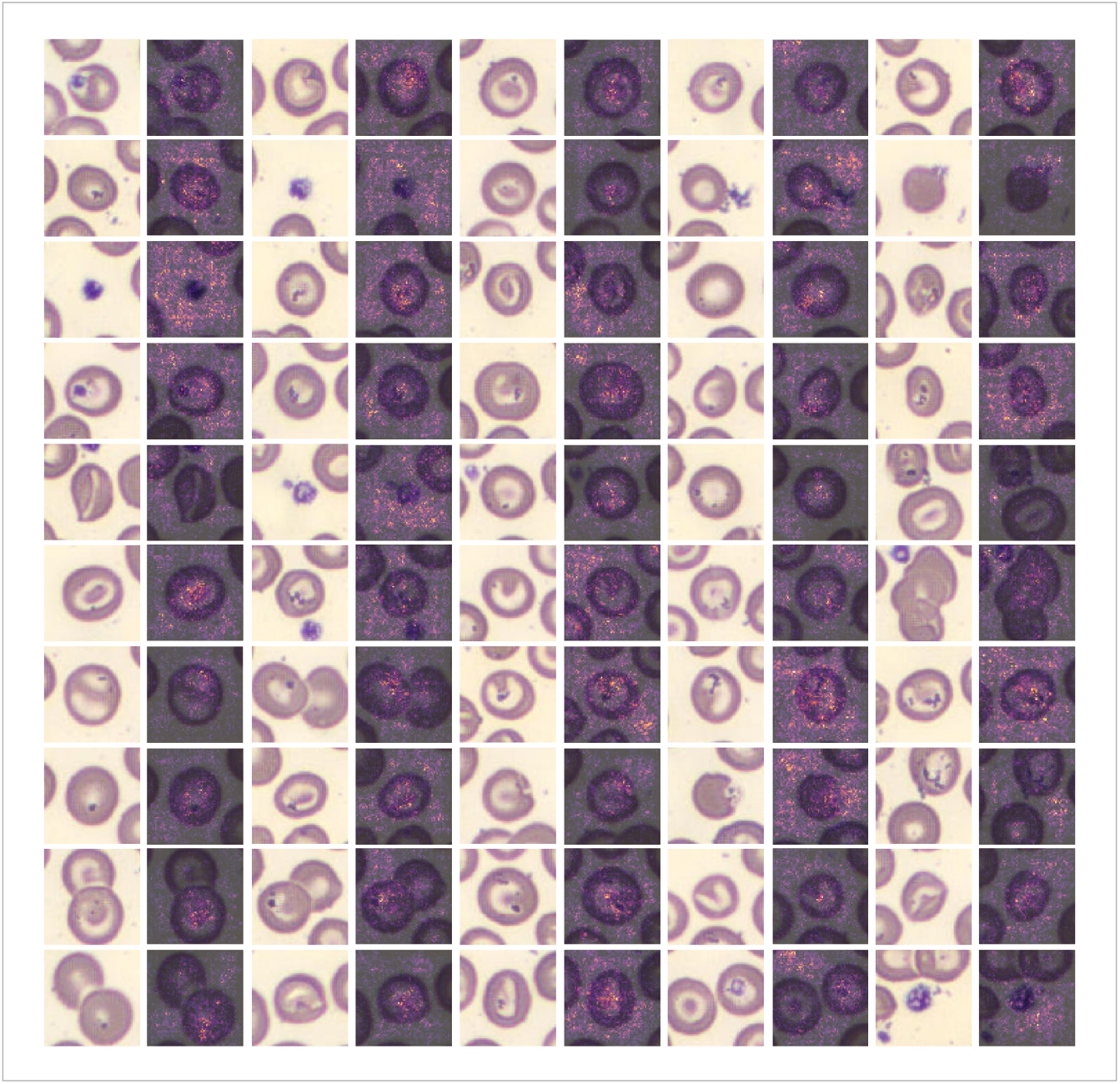
Integrated gradient (IG) visualizations including the original image and an overlay of the pixel-wise IG attribution mask and the original image. Images are from Case #25 of the clinical validation dataset and are those which were predicted as belonging to the ‘parasite’ class.

## DISCUSSION

In this report, we describe an approach to quantifying percent-parasitemia in peripheral blood smears using computer vision and machine learning technology. We sought to examine the accuracy of an ML-based solution without the use of expert operator-reclassification. Since the beginning of modern computing, there has been considerable interest in the optimization of peripheral blood smear review, with published efforts for smear image analysis dating back to the 1970’s (14,15). While previous attempts yielded variable results, recent improvements in computing hardware have led to significant advancements in performance, particularly in the context of object classification tasks (16). Indeed, there has been a resurgence over recent years investigating the application of machine learning-based technologies for classification, speciation, and quantitative tasks using digital images of the peripheral blood smear (17,18). Automated image analysis tools are becoming increasingly available for peripheral smear analysis, however, the scope of FDA approval is limited and classification algorithms demonstrate suboptimal performance without human reclassification (19,20).

We found that in the context of the train-test development cycle, model performance metrics demonstrated highly accurate results. Train and validation loss curves demonstrated minimally appreciable divergence towards the end of training iterations which would imply that there is negligible overfitting with the cell classification model (Figure 3A). The sigmoid activation function used for the classification layer of the model demonstrated good separation between the parasite class and the non-parasite class, with only 20 false positive cells in the test dataset (Figure 4). However, when the model was implemented with contour-based cell segmentation and applied to the clinical validation dataset, method comparison studies with the MLS-RS demonstrated suboptimal concordance with the model-based method. Simple linear regression between the two methods had a calculated correlation coefficient (R2) of 0.244 and 0.916 with and without outliers, respectively. In addition, only 7 of the 14 positive cases were concordant between the model and MLS-RS when grouped by percent parasitemia bins. Lastly, there were three major errors by the model-based method, which were defined as discordance around the clinical decision point of 10% parasitemia (Supplemental Table 2).

The root cause of these discrepancies is likely multifactorial and highlights the need to interrogate the performance of ML-based technology beyond the train-test development cycle. In the clinical validation method-to-method comparison, the model returned highly erroneous positive signal with cases 11 and 16, relative to the MLS-RS (Figure 5). These errors were likely driven, in part, by the quality of the blood smear which contained significant amount of precipitate and rouleaux formations. For blood smear images where there was minimal to no rouleaux formation, visual inspection of contour-based cell segmentation suggested adequate performance (Supplemental Figure 2). However, in the context of significant rouleaux formation, cell segmentation resulted in fewer individual cells identified for evaluation (Supplemental Figure 3 and 4). In combination with overlying precipitate, which can be mistaken for intra-erythrocyte parasites, this can result in a high numerator (i.e., false positives) and a low denominator (i.e., fewer individually segmented cells), which led to artificially elevated parasitemia quantification. Future work in this area could explore the use of ML-based approaches to cell segmentation. However, these approaches would theoretically encounter similar barriers when initializing models with coordinates for segmentation training and would need specific considerations for handling rouleaux formations. During the initial stages of this work, we had found there to be little qualitative difference between computer vision and ML-based segmentation for smears when there was minimal rouleaux formation to contend with (data not shown).

Model interpretability experiments were used to develop an intuitive sense as to what effectuates the observed model behavior, a limitation being that this method only provides an indication of feature importance on individual images and does not offer a mechanism to provide insight across the entire dataset. It also only explains individual feature contributions, but does not examine how feature interactions may contribute to predictions (21). Nonetheless, these experiments revealed that model predictions of the target class, ‘parasite’, were generally most impacted by pixels spatially related to intraerythrocytic ring-forms (Figure 7). However, there were instances wherein pixel-wise activation patterns were found to be localized outside of the erythrocyte and corresponding to background noise (Supplemental Figure 5). This would suggest that there is some degree of overfitting which is not obviously appreciable through visual inspection of the train and validation loss curves. Integrated gradients also provided some context as to model fallibility when applied to the clinical validation dataset. Cells which were classified as ‘parasite’ from case 25 demonstrated pixel-wise activation patterns which suggest that the model prediction of the target class was susceptible to influence by features which share similarities to ring-form parasites. Examples of these microscopic features which were associated with localized pixel activation included variations in erythrocyte morphology (e.g., target cell contours) and overlying precipitate or platelets (Figure 8).

In general, model misclassification errors may be remedied by increasing the number of class examples during training. In doing so, the model input space would be more representative of the heterogeneity the model may be expected to encounter with real-world data, relative to a model trained with fewer class examples. However, in the context of training classification models in healthcare, particularly those which rely on cases of low prevalence diseases, increasing the number of training examples can be prohibitive. There are techniques which can be implemented to artificially expand the size of the training dataset (e.g., label-preserving image transformations) and improve model performance and generalizability. However, these techniques are limited in terms of their performance benefits and cannot portray inherent intra-class variability which is not already represented in the existing training dataset. Overall, results of this study reinforce the need for consistent, artifact-free, high quality data for optimal algorithm performance.

Most scientific literature on parasite quantitation is done in the context of Malaria diagnostics, whereas approaches leveraging deep learning methods have only recently been described (22). To our knowledge, this is the first published work to focus on the quantitation of *Babesia* with interpretable clinical results, using images that are derived from routine clinical workflows. Further, we also evaluated the utility of the model-based method using external validation datasets, not commonly done in malaria quantitation studies (18,23). Similar to other published reports, we classified and quantified intracellular parasites using ‘per-cell’ images (24). Other articles have also described a region-based approach, wherein images containing multiples cells are evaluated for intracellular parasites, and a final quantitative score is ultimately produced (18). However, while there are arguably benefits to each, there is currently no clear advantage to either approach. Indeed, with the increasing breadth of machine learning technologies, there are multiple avenues to pursue for parasite quantitation. Further research is needed to delineate which methods are most performant, scalable, and most easily implemented into clinical workflows, as well as addressing data quality for machine learning implementation in microscopic image-based computer analysis.

## Data Availability

Data will not be made publicly available.

## ACKNOWLEDGEMENTS

We would like to acknowledge Lisa Mehlin, Holly Base, and Laura Pires for volunteering their time to quantitate *Babesia* parasites for the purposes of the clinical validation portion of this study. We would also like to acknowledge John Errico and Cai Mayberry for their administrative support of this work.

## Abbreviations

AUC: Area Under the Curve
FN: False Negative
FP: False Positive
IG: Integrated Gradient
MLS: Medical Laboratory Scientist
MLS-RS: Medical Laboratory Scientist-Reference Standard
RBC: Red blood cell
TN: True Negative
TP: True Positive
XAI: Explainable Artificial Intelligence

