## Supplemental Material for "Applications of Digital Microscopy and Densely Connected Convolutional Neural Networks for Automated Quantitation of Babesia-Infected Erythrocytes"

**Supplemental Table 1**: Summary statistics of clinical reference standard measurements. ‘Total’ represents the average, SD, and CV, across all MLS reads (n=9).

| | MLS-A | | | MLS-B | | | MLS-C | | | Total | | |
|---|---|---|---|---|---|---|---|---|---|---|---|---|
| Case Number | Average | SD | CV | Average | SD | CV | Average | SD | CV | Average | SD | CV |
| 1 | 39.43 | 2.36 | 5.99 | 32.90 | 0.91 | 2.76 | 43.17 | 12.32 | 28.55 | 38.50 | 8.41 | 21.85 |
| 2 | 19.03 | 0.37 | 1.93 | 15.23 | 0.42 | 2.75 | 16.27 | 1.89 | 11.60 | 16.84 | 1.97 | 11.67 |
| 3 | 20.63 | 4.78 | 23.14 | 13.27 | 0.97 | 7.29 | 15.20 | 0.75 | 4.92 | 16.37 | 4.22 | 25.80 |
| 4 | 13.37 | 1.09 | 8.13 | 10.73 | 2.10 | 19.55 | 11.73 | 1.72 | 14.67 | 11.94 | 2.01 | 16.80 |
| 5 | 11.60 | 2.41 | 20.74 | 8.63 | 1.13 | 13.12 | 9.70 | 2.94 | 30.35 | 9.98 | 2.60 | 26.04 |
| 6 | 9.93 | 1.30 | 13.11 | 8.30 | 1.14 | 13.77 | 9.43 | 1.01 | 10.75 | 9.22 | 1.35 | 14.59 |
| 7 | 10.07 | 1.72 | 17.10 | 4.47 | 0.37 | 8.24 | 5.00 | 0.86 | 17.28 | 6.51 | 2.77 | 42.48 |
| 8 | 5.17 | 0.25 | 4.83 | 3.77 | 0.91 | 24.17 | 3.80 | 1.24 | 32.52 | 4.24 | 1.11 | 26.14 |
| 9 | 3.63 | 0.24 | 6.49 | 2.63 | 0.54 | 20.64 | 3.10 | 0.59 | 18.99 | 3.12 | 0.63 | 20.24 |
| 10 | 3.23 | 0.49 | 15.22 | 1.90 | 0.29 | 15.49 | 1.97 | 0.09 | 4.79 | 2.37 | 0.70 | 29.54 |
| 11 | 2.80 | 1.31 | 46.93 | 2.27 | 0.17 | 7.50 | 1.57 | 0.61 | 39.12 | 2.21 | 0.98 | 44.44 |
| 12 | 2.37 | 0.87 | 36.89 | 2.00 | 0.50 | 24.83 | 2.00 | 0.49 | 24.49 | 2.12 | 0.67 | 31.47 |
| 13 | 1.63 | 0.63 | 38.83 | 0.87 | 0.17 | 19.61 | 0.83 | 0.31 | 37.09 | 1.11 | 0.56 | 50.28 |
| 14 | 0.70 | 0.22 | 30.86 | 0.37 | 0.05 | 12.86 | 0.37 | 0.12 | 34.02 | 0.48 | 0.21 | 44.97 |
| 16 | 0.00 | 0.00 | 0.00 | 0.00 | 0.00 | 0.00 | 0.00 | 0.00 | 0.00 | 0.00 | 0.00 | 0.00 |
| 17 | 0.00 | 0.00 | 0.00 | 0.00 | 0.00 | 0.00 | 0.00 | 0.00 | 0.00 | 0.00 | 0.00 | 0.00 |
| 18 | 0.00 | 0.00 | 0.00 | 0.00 | 0.00 | 0.00 | 0.00 | 0.00 | 0.00 | 0.00 | 0.00 | 0.00 |
| 19 | 0.00 | 0.00 | 0.00 | 0.00 | 0.00 | 0.00 | 0.00 | 0.00 | 0.00 | 0.00 | 0.00 | 0.00 |
| 20 | 0.00 | 0.00 | 0.00 | 0.00 | 0.00 | 0.00 | 0.00 | 0.00 | 0.00 | 0.00 | 0.00 | 0.00 |
| 21 | 0.00 | 0.00 | 0.00 | 0.00 | 0.00 | 0.00 | 0.00 | 0.00 | 0.00 | 0.00 | 0.00 | 0.00 |
| 22 | 0.00 | 0.00 | 0.00 | 0.00 | 0.00 | 0.00 | 0.00 | 0.00 | 0.00 | 0.00 | 0.00 | 0.00 |
| 23 | 0.00 | 0.00 | 0.00 | 0.00 | 0.00 | 0.00 | 0.00 | 0.00 | 0.00 | 0.00 | 0.00 | 0.00 |
| 24 | 0.00 | 0.00 | 0.00 | 0.00 | 0.00 | 0.00 | 0.00 | 0.00 | 0.00 | 0.00 | 0.00 | 0.00 |
| 25 | 0.00 | 0.00 | 0.00 | 0.00 | 0.00 | 0.00 | 0.00 | 0.00 | 0.00 | 0.00 | 0.00 | 0.00 |

CV = Coefficient of Variation; SD = Standard Deviation

**Supplemental Figure 1**: Summary statistics of clinical reference standard measurements. Bars represent average of parasitemia measurements per MLS (n=3 per MLS). Error bars represent 1 standard deviation.

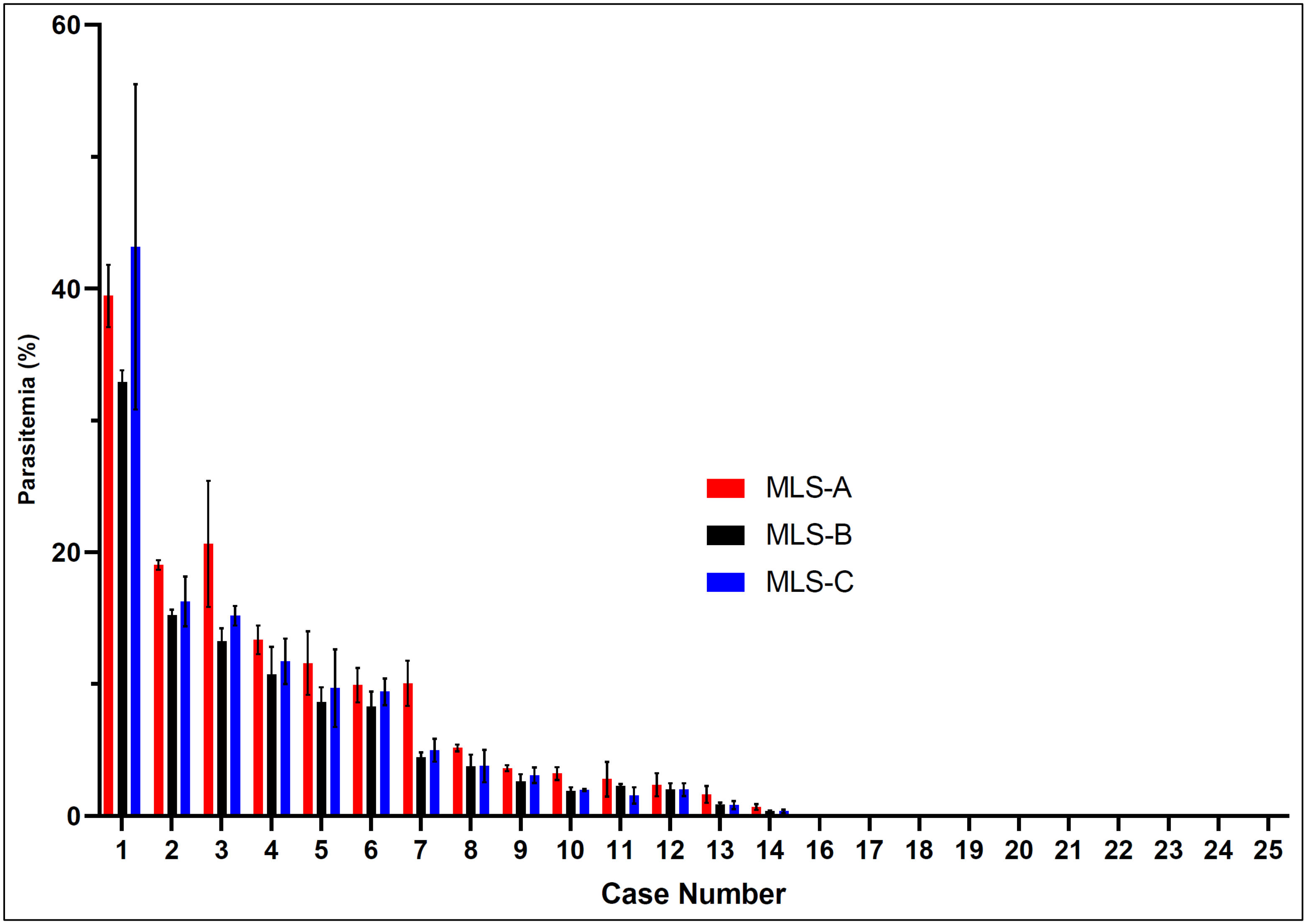

**Supplemental Table 2**: Table of mean (SD) parasitemia result values for clinical validation cases. Cells are highlighted to denote categorically defined major (red) and minor (yellow) errors. Major errors are defined as discrepancy between the model and the MLS-reference standard around the clinical decision point of 10% parasitemia. Minor errors are defined as discrepancy between CAP PSR-based categories.

| Case Number | MLS Mean (SD) | Model Mean (SD) | MLS CV | Model CV | MLS Category | Model Category |
|---|---|---|---|---|---|---|
| 1 | 38.5 (8.9) | 27.2 (1) | 23.2 | 3.6 | >9.9 | >9.9 |
| 2 | 16.8 (2.1) | 17.9 (2.9) | 12.4 | 16.1 | >9.9 | >9.9 |
| 3 | 16.4 (4.5) | 12.2 (1.8) | 27.4 | 14.6 | >9.9 | >9.9 |
| 4 | 11.9 (2.1) | 8.6 (1.1) | 17.8 | 13.3 | >9.9 | 5.0-9.9 |
| 5 | 10 (2.8) | 6 (0.3) | 27.6 | 4.6 | >9.9 | 5.0-9.9 |
| 6 | 9.2 (1.4) | 6.9 (0.2) | 15.5 | 3.3 | 5.0-9.9 | 5.0-9.9 |
| 7 | 6.5 (2.9) | 3.8 (0.3) | 45.1 | 9.0 | 5.0-9.9 | 2.0-4.9 |
| 8 | 4.2 (1.2) | 4.7 (0.7) | 27.7 | 15.5 | 2.0-4.9 | 2.0-4.9 |
| 9 | 3.1 (0.7) | 5 (1)¥ | 21.5 | 21.1 | 2.0-4.9 | 5.0-9.9 |
| 10 | 2.4 (0.7) | 8.4 (0.4) ¥ | 31.3 | 4.3 | 2.0-4.9 | 5.0-9.9 |
| 11 | 2.2 (1) | 39.2 (19.1) ¥ | 47.1 | 48.7 | 2.0-4.9 | >9.9 |
| 12 | 2.1 (0.7) | 2.9 (0.1) | 33.4 | 4.3 | 2.0-4.9 | 2.0-4.9 |
| 13 | 1.1 (0.6) | 3 (0.6) ¥ | 53.3 | 20.0 | 1.0-1.9 | 2.0-4.9 |
| 14 | 0.5 (0.2) | 0.3 (0.1) | 47.7 | 25.5 | 0.1-0.9 | 0.1-0.9 |
| 16 | 0 (0) | 24.1 (21) ¥ | | 87.2 | 0.0 | >9.9 |
| 17 | 0 (0) | 0.4 (0.3) ¥ | | 78.9 | 0.0 | 0.1-0.9 |
| 18 | 0 (0) | 1.5 (0.5) ¥ | | 33.3 | 0.0 | 1.0-1.9 |
| 19 | 0 (0) | 1.9 (0.3) ¥ | | 15.3 | 0.0 | 1.0-1.9 |
| 20 | 0 (0) | 1.7 (0.3) ¥ | | 15.3 | 0.0 | 1.0-1.9 |
| 21 | 0 (0) | 0.5 (0.2) ¥ | | 47.0 | 0.0 | 0.1-0.9 |
| 22 | 0 (0) | 1 (0.6) ¥ | | 55.6 | 0.0 | 1.0-1.9 |
| 23 | 0 (0) | 1.3 (0) ¥ | | 0.0 | 0.0 | 1.0-1.9 |
| 24 | 0 (0) | 1.8 (0.6) ¥ | | 35.8 | 0.0 | 1.0-1.9 |
| 25 | 0 (0) | 3.4 (0.1) ¥ | | 3.2 | 0.0 | 2.0-4.9 |

¥ = Model mean is more than 2 SD from MLS mean

**Supplemental Figure 2:** Representative image of contour-based erythrocyte segmentation.

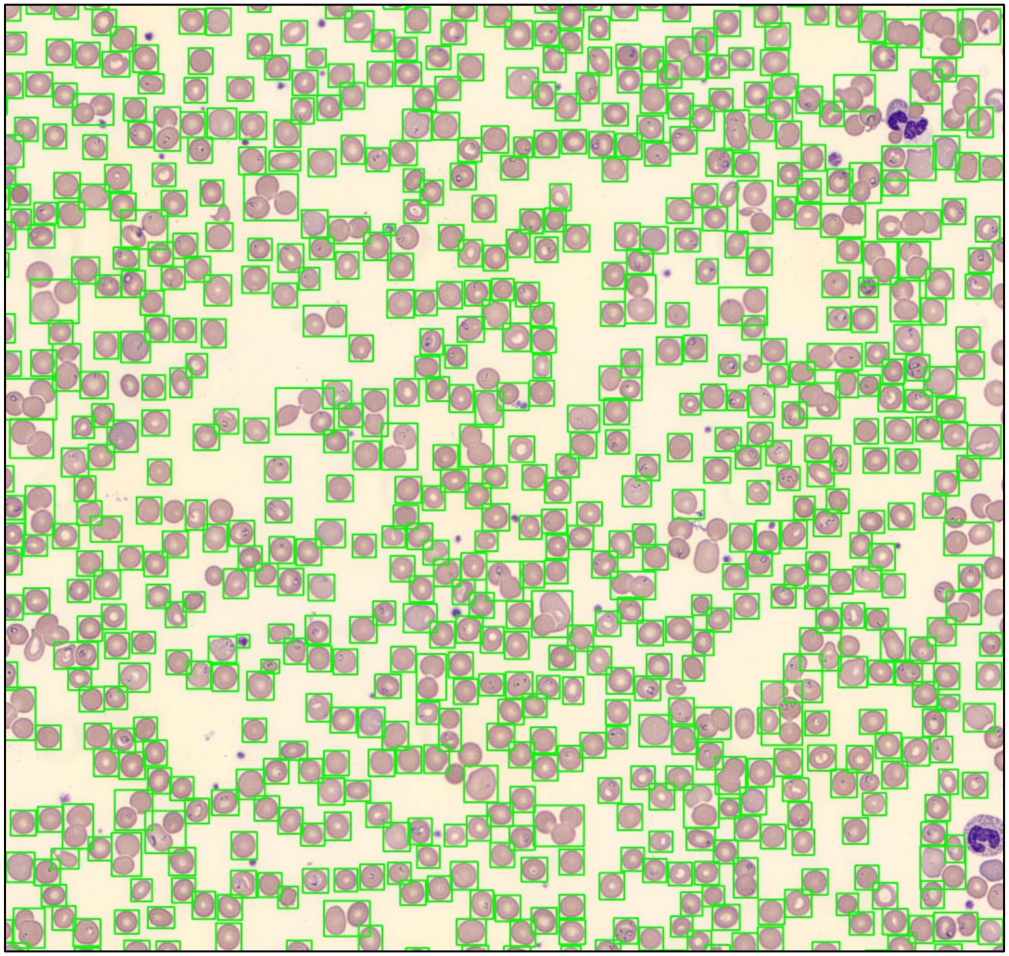

**Supplemental Figure 3**: Representative image of Case #11

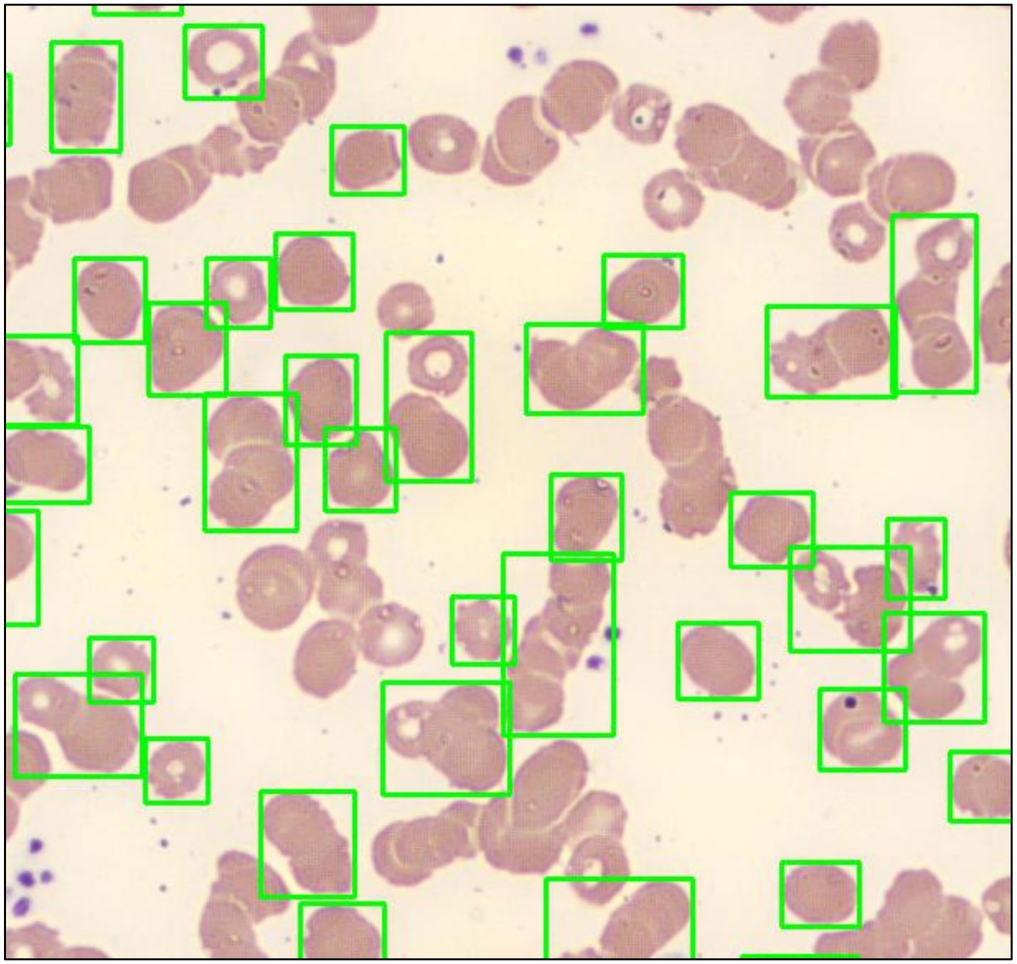

**Supplemental Figure 4**: Representative image of Case #16 Image #1

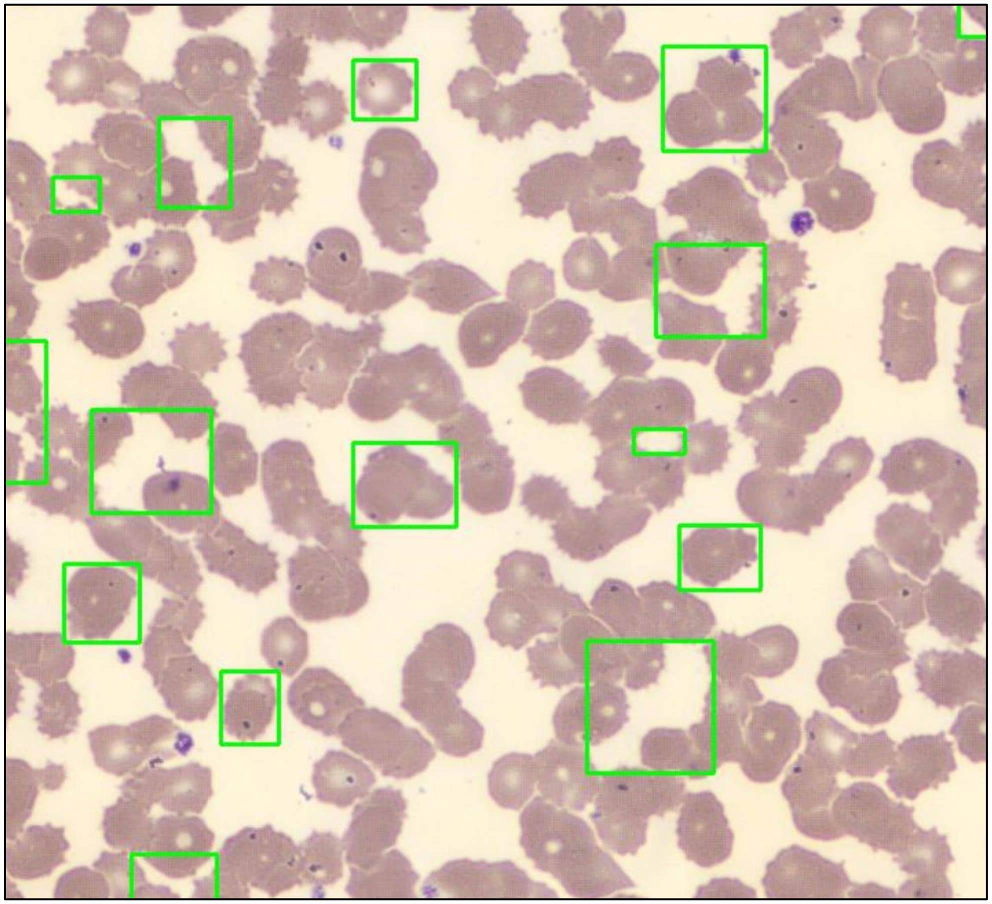

**Supplemental Figure 5**: Representative example of a predicted class of 'parasite' with integrated gradient derived pixel-wise activation patterns shown, and not localized to the intra-erythrocytic ring-form.

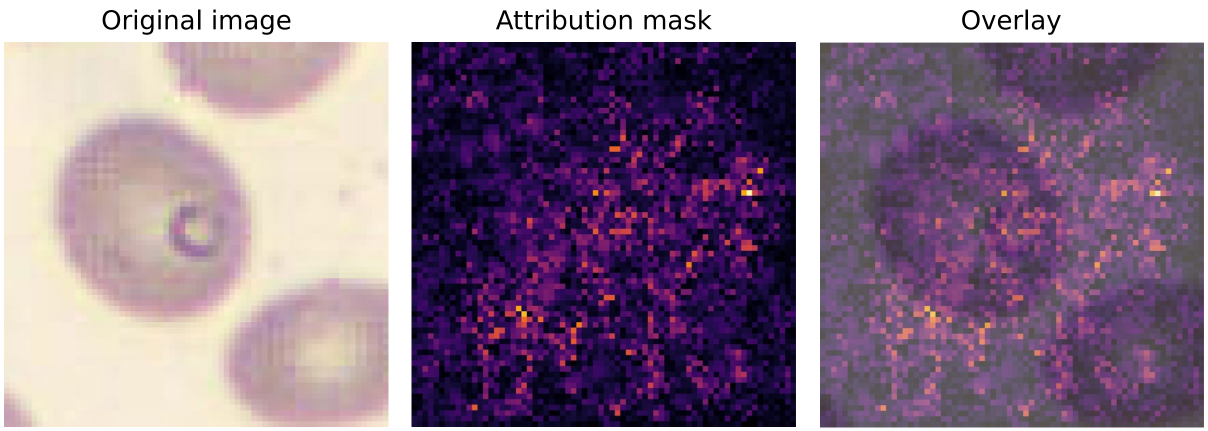
